# Performance of Existing and Novel Symptom- and Antigen Testing-Based COVID-19 Case Definitions in a Community Setting

**DOI:** 10.1101/2022.05.10.22274914

**Authors:** Scott Lee, Olivia Almendares, Jessica L. Prince-Guerra, Charles M. Heilig, Jacqueline E. Tate, Hannah L. Kirking

## Abstract

Point-of-care antigen tests are an important tool for SARS-CoV-2 detection, but they are less clinically sensitive than real-time reverse-transcription PCR (RT-PCR), impacting their efficacy as screening procedures. Our goal in this study was to see whether we could improve this sensitivity by considering antigen test results in combination with other relevant information, namely exposure status and reported symptoms. In November of 2020, we collected 3,419 paired upper respiratory specimens tested by RT-PCR and the Abbott BinaxNOW antigen test at two community testing sites in Pima County, Arizona. We used symptom, exposure, and antigen testing data to evaluate the sensitivity and specificity of various symptom definitions in predicting RT-PCR positivity. Our analysis yielded 6 novel multi-symptom case definitions with and without antigen test results, the best of which overall achieved a Youden’s J index of 0.66, as compared with 0.52 for antigen testing alone. Using a random forest as a guide, we show that this definition, along with our others, does not lose the ability to generalize well to new data despite achieving optimal performance in our sample. Our methodology is broadly applicable, and we have made our code publicly available to aid public health practitioners in developing or fine-tuning their own screening rules.

## Introduction

Since the emergence of severe acute respiratory syndrome coronavirus 2 (SARS-CoV-2) in December 2019, the virus has spread globally, causing widespread illness with varying degrees of severity. The emergence of several SARS-CoV-2 variants over time has led to periodic spikes in cases, with the most recent surge in cases related to the B.1.1.529 variant (Omicron) . Testing is an important part of the overall strategy to reduce and/or control SARS-CoV-2 spread, and antigen testing in particular has become very common, with tests widely available for both clinical and at-home use. Relative to nucleic acid amplification tests (NAATs) for SARS-CoV2, antigen tests have several benefits: operationally, they are fast, cheap, and relatively easy to administer; and diagnostically, they are very specific, with false positive rates often under 1%. However, the clinical sensitivity of these tests varies substantially, depending, among other things, on the patient’s exposure status, severity of illness, duration of illness, and presence of symptoms (2-8). For the Abbott BinaxNOW COVID-19 Ag Card (BinaxNOW) antigen test, which has been widely adopted in the United States for both community screening and at-home testing, investigators have found sensitivities ranging from as low as approximately 50% (3, 5, 9-12) to as high as 92% (11), with many more in between (e.g. from 74%-77% (6, 7, 13, 14)). These values are much lower than the test’s reported sensitivity of 97%, and their variability makes it difficult for public health practitioners to estimate accurately the performance of their screening programs without relying on routine confirmatory testing with NAATs.

When potential COVID-19 cases are asymptomatic, antigen testing is more feasible and perhaps the better screening option, particularly when epidemiologic risk factors for infection (such as travel history or close contact with a confirmed case) have not been identified. When potential cases are symptomatic, however, the screening scenario changes in two important ways: the sensitivity of the antigen testing tends to improve (Prince-Guerra 2021), and there is more clinical information available for predicting the underlying infection. The current Council of State and Territorial Epidemiologists (CSTE) COVID-19 surveillance case definition (15) follows this line of reasoning for identifying probable cases, requiring them to have either a positive antigen test or to have had close contact with a confirmed case and be symptomatic. To the best of our knowledge, this case definition was not directly derived from case-level clinical and laboratory data (e.g., using the kind of optimization procedure we adopt below), meaning it may not provide an optimal balance of sensitivity and specificity. What balance it does provide is fixed, since it is a single binary rule intended for surveillance, rather than a collection of rules with different operational characteristics intended for a variety of clinical use cases. The question remains, then, of how best to combine symptom information with antigen test results for screening.

In this analysis, our goal was to show how this question might be answered by rigorously exploring the sensitivity-specificity tradeoff in these sorts of hybrid, testing/symptom-based screening case definitions. Building on previous work in improving screening and diagnosis procedures for tuberculosis disease (16) and COVID-19 (17), we show that a brute-force combinatorial search, along with some guidance from a robust machine learning model, can find definitions that improve the sensitivity of antigen testing relative to RT-PCR without sacrificing too much of its specificity. Although the exact definitions we propose may not be appropriate for all COVID-19 screening or diagnosis applications, the discovery procedure itself is adaptable to any dataset with symptom data and a reference standard (e.g., RT-PCR) and will thus be useful for both researchers and practitioners alike in developing definitions tailored to their own scientific and programmatic uses.

## Methods

### Data collection

We collected paired upper respiratory swabs, demographics, symptom information and exposure history from participants aged 10 years and older at two community testing sites in Pima County, Arizona during November 3-17, 2020, as previously described (3, 10). Here we use symptom data which captured presence of 15 symptoms in the 14 days prior to or on the day of testing. The list of symptoms includes all symptoms per CSTE COVID-19 2020 Interim Case Definition (15). The protocol for this evaluation was reviewed by the Centers for Disease Control and Prevention (CDC) and determined to be non-research and was conducted consistent with applicable federal law and CDC policy as defined in 45 CRF 46.102(I)(2) (18).

All paired swab specimens were tested by RT-PCR and the Abbott BinaxNOW COVID-19 Ag Card (BinaxNOW) antigen test. A healthcare professional collected a bilateral anterior nasal swab first for BinaxNOW testing, immediately followed by a bilateral nasopharyngeal swab for RT-PCR testing. Nasopharyngeal swabs were tested using either the CDC 2019-nCoV RT-PCR Diagnostic Panel (CDC RT-PCR assay) for detection of SARS-CoV-2 (n=2,582) (19) or the Fosun COVID-19 RT-PCR Detection Kit (Fosun RT-PCR assay) (n=837) (20). As described previously (3), validation by the commercial laboratory reported the limit of detection as equivalent at 500 copies/mL for both assays. In the analysis below, both tests were treated equally as reference standards for classification.

### Analytic aims

Our main goal was to see whether we could improve the performance of antigen testing in predicting RT-PCR positivity by using a hybrid case definition that also considers reported symptoms. To that end, we explored two methods for arriving at such a case definition: heuristics based on combinations of reported symptoms and antigen test results, including an exhaustive combinatorial search of the various predictors; and heuristics based on a tree-based machine learning model trained on the data. We were also interested in determining which of the hybrid case definitions would be most likely to generalize out-of-sample data. Here, we used two heuristics to avoid this kind of overfitting: a preference for parsimony, such that case definitions considering fewer variables are preferable to ones considering more; and a preference for case definitions that do not grossly exceed the generalization performance of the same machine learning model as above.

### Heuristics based on random forests

To estimate the maximum generalizable classification performance, we might expect from these hybrid case definitions, we turned to the random forest (21), a tree-based machine learning model theoretically guaranteed not to overfit its training data. We trained random forests on two feature sets: one using only the individual symptoms as predictors, and one using both the individual symptoms and antigen test results as predictors. For each set of predictors, we fit five large forests (each with 10,000 trees) to the entire dataset, beginning with trees of depth one for the first forest, and increasing the depth of the trees by one for each successive forest. This process yielded 10 trained forests, one for each unique combination of feature set (n=2) and depth (n=5).

From each of these 10 forests, we retrieved the out-of-bag decision scores for all observations, yielding a forest-derived distribution of predicted probabilities for RT-PCR positivity. We then compared the predicted probabilities with the actual RT-PCR test results to construct a standard receiver operating characteristic (ROC) curve, plotting sensitivity as a function of 1 - specificity. Because this curve contains the random forest’s best-possible scores for Youden’s J index, we treated it as an approximate upper bound on the generalization performance we might expect from naive (i.e., non-learned) case definitions constructed from the same set of variables.

In addition to estimating the upper bound on generalizable classification performance, we also used the random forest as a heuristic for selecting symptoms to include in a combined case definition with the antigen results. After training, random forests rank their features based on the average amount of information they contributed to the constituent trees’ predictions. Here, we used the feature importances from the symptoms-and-antigen forests to determine which symptoms worked best in combination with test results. We note here that the decision trees in a random forest may split on features “both ways”, such that an important feature may be a lack of that feature and not a presence.

### Heuristics based on combinatorial analysis

We used a brute-force combinatorial approach to enumerate the full range of classification scores achievable with non-machine-learned case definitions. Here, we considered three sets of predictors: the symptoms alone, the symptoms in combination with antigen test results, and the symptoms plus known exposure to a confirmed case in combination with the antigen test results. For the symptoms alone, we enumerated all possible *m*-of-*n* combinations of symptoms, where *n* is the total number of symptoms in the combination, and *m* is the minimum number of symptoms required to be present for the patient to be considered a case. We used 5 as the maximum value for both *m* and *n*, mainly to limit the computational complexity of the enumeration, but also to improve the generalizability of the results. For the symptoms and antigen testing together, we combined this enumeration with the binary antigen test results, such that anyone with either a positive antigen test or *m* of the *n* symptoms in a given combination would be labeled a case.

### Established case definitions

To serve as a point of comparison for the combinations, we evaluated the predictive performance of 3 previously established case definitions: the CSTE COVID-19 case definition (15), and two compound symptom combinations found to work well in a published development study (17). We redefine these last two as follows:

*Reses 1*: loss of taste or smell OR at least 1 of (shortness of breath, fever and/or chills, or myalgia)

*Reses 2*: loss of taste or smell OR at least 3 of (shortness of breath, difficulty breathing, fever and/or chills, or myalgia)

Both these definitions worked well in the development study; *Reses 2* achieved the highest overall performance as measured by F1 score (80%). To our knowledge, this is the first time the definitions have been validated on external data, as well as having their performance examined in combination with antigen test results.

### Classification metrics

For the purposes of screening and diagnosis in the absence of confirmatory testing, we evaluated prediction methods on several common measures of binary classification performance, including sensitivity (se), specificity (sp), Youden’s J index, and F1 score. For surveillance purposes, we evaluated methods on a single metric, the difference between actual prevalence, defined as the number of RT-PCR positive patients in our dataset, and predicted prevalence, defined as the number of positive calls from the classification method.

### Statistical Analysis

For inference, we constructed bias-corrected and accelerated bootstrap confidence intervals (22) for all metrics; these were not adjusted for multiplicity. We also used the Wilcoxon signed-rank test to differences in paired classification metrics, where appropriate.

### Software and hardware

The random forest, combinatorial search, and statistical analysis were implemented in Python 3.8 with the scikit-learn (23) and statsmodels (24) packages. We ran all analyses on a scientific workstation with a 12-core CPU and 128 GB of RAM, and we used parallelization whenever possible to improve runtime. Our code is publicly available on GitHub at https://github.com/scotthlee/az-covid.

## Results

We analyzed 3,419 paired specimens from 3,302 total participants aged 10 years and older with complete testing and survey data for which demographic and symptom data has been previously described in detail (3).

### Performance of single predictors

Table 1 shows the performance of the single symptoms and antigen test results in predicting RT-PCR positivity. In terms of individual-level performance, congestion was the most sensitive (62%), followed by headache (59%), antigen testing (52%), cough (38%), and chills (37%). Antigen testing was the most specific (99%) and was closely followed by loss of taste or smell (97%), difficulty breathing (97%), rigors (shivers) (96%), and abdominal pain (96%). Overall, antigen testing was the best single predictor, with the highest Youden’s J index (52%) and F1- score (68%), even though its sensitivity was relatively low (53%). In terms of population-level performance, chills had the most accurate prevalence estimate (+4%), followed by diarrhea (−12%) and fever (−20%).

**Table 1.** Point estimates and 95% confidence intervals for the classification performance of single-symptom case definitions and for antigen testing.

|  | Sensitivity<sup>1</sup> | Specificity<sup>2</sup> | PPV<sup>3</sup> | NPV<sup>4</sup> | Youden's J<sup>5</sup> | F1 score<sup>6</sup> | Relative Difference in Prevalence<sup>7</sup> |
| --- | --- | --- | --- | --- | --- | --- | --- |
|  | Value (95% CI) | Value (95% CI) | Value (95% CI) | Value (95% CI) | Value (95% CI) | Value (95% CI) | Value (95% CI) |
| **Symptoms** |  |  |  |  |  |  |  |
| Fever<sup>8</sup> | 0.3 (0.27, 0.36) | 0.95 (0.94, 0.96) | 0.38 (0.34, 0.44) | 0.93 (0.92, 0.94) | 0.26 (0.22, 0.32) | 0.34 (0.3, 0.39) | -0.2 (-0.29, -0.1) |
| Chills | 0.37 (0.31, 0.42) | 0.94 (0.92, 0.94) | 0.35 (0.3, 0.4) | 0.94 (0.93, 0.94) | 0.3 (0.25, 0.35) | 0.36 (0.31, 0.4) | 0.04 (-0.08, 0.18) |
| Shivers/Rigors | 0.21 (0.17, 0.25) | 0.96 (0.96, 0.97) | 0.37 (0.31, 0.45) | 0.93 (0.92, 0.93) | 0.18 (0.14, 0.23) | 0.27 (0.22, 0.33) | -0.42 (-0.51, -0.33) |
| Muscle Aches | 0.47 (0.42, 0.53) | 0.89 (0.88, 0.9) | 0.29 (0.24, 0.32) | 0.95 (0.94, 0.95) | 0.36 (0.3, 0.42) | 0.36 (0.31, 0.4) | 0.63 (0.45, 0.82) |
| Congestion<sup>9</sup> | 0.62 (0.58, 0.68) | 0.76 (0.75, 0.78) | 0.2 (0.18, 0.23) | 0.95 (0.95, 0.96) | 0.38 (0.34, 0.45) | 0.3 (0.28, 0.34) | 2.07 (1.71, 2.41) |
| Sore Throat | 0.41 (0.35, 0.47) | 0.85 (0.84, 0.87) | 0.21 (0.18, 0.25) | 0.94 (0.93, 0.94) | 0.27 (0.2, 0.32) | 0.28 (0.25, 0.33) | 0.97 (0.74, 1.14) |
| Cough<sup>10</sup> | 0.38 (0.34, 0.44) | 0.9 (0.89, 0.91) | 0.27 (0.24, 0.32) | 0.94 (0.93, 0.94) | 0.28 (0.24, 0.34) | 0.32 (0.28, 0.36) | 0.41 (0.26, 0.57) |
| Shortness of Breath<sup>10</sup> | 0.17 (0.13, 0.21) | 0.95 (0.94, 0.96) | 0.24 (0.18, 0.3) | 0.92 (0.91, 0.93) | 0.12 (0.08, 0.16) | 0.2 (0.15, 0.25) | -0.31 (-0.41, -0.2) |
| Difficulty breathing | 0.1 (0.07, 0.13) | 0.97 (0.96, 0.98) | 0.24 (0.18, 0.33) | 0.92 (0.91, 0.93) | 0.07 (0.04, 0.11) | 0.14 (0.11, 0.19) | -0.58 (-0.7, -0.54) |
| Nausea or vomiting | 0.15 (0.11, 0.19) | 0.94 (0.94, 0.95) | 0.21 (0.16, 0.26) | 0.92 (0.91, 0.93) | 0.1 (0.06, 0.13) | 0.17 (0.14, 0.21) | -0.27 (-0.37, -0.17) |
| Headache | 0.59 (0.55, 0.64) | 0.8 (0.78, 0.81) | 0.22 (0.2, 0.25) | 0.95 (0.94, 0.96) | 0.39 (0.36, 0.46) | 0.32 (0.29, 0.35) | 1.72 (1.46, 1.98) |
| Abdominal Pain | 0.09 (0.07, 0.15) | 0.96 (0.95, 0.96) | 0.18 (0.13, 0.25) | 0.92 (0.9, 0.92) | 0.05 (0.03, 0.09) | 0.12 (0.09, 0.19) | -0.49 (-0.55, -0.37) |
| Diarrhea | 0.17 (0.14, 0.21) | 0.93 (0.92, 0.94) | 0.19 (0.15, 0.24) | 0.92 (0.91, 0.93) | 0.1 (0.07, 0.14) | 0.18 (0.15, 0.22) | -0.12 (-0.24, 0) |
| Loss of Taste or Smell | 0.3 (0.26, 0.35) | 0.97 (0.97, 0.98) | 0.53 (0.48, 0.59) | 0.94 (0.93, 0.94) | 0.28 (0.23, 0.32) | 0.39 (0.34, 0.44) | -0.42 (-0.51, -0.35) |
| Fatigue | 0.44 (0.4, 0.49) | 0.84 (0.83, 0.85) | 0.21 (0.19, 0.24) | 0.94 (0.93, 0.95) | 0.29 (0.24, 0.34) | 0.29 (0.26, 0.32) | 1.07 (0.86, 1.35) |
| **Antigen** | 0.53 (0.47, 0.58) | 0.99 (0.99, 1.0) | 0.98 (0.95, 0.99) | 0.95 (.95, .96) | 0.52 (0.47, 0.58) | 0.68 (0.64, 0.73) | -0.46 (-0.52, -0.39) |

<sup>1</sup> Sensitivity, the probability of testing positive when disease is present.
<sup>2</sup> Specificity, the probability of testing negative when disease is absent.
<sup>3</sup> Positive predictive value, the probability of a patient having disease when test is positive.
<sup>4</sup> Negative predictive value, the probability of a patient not having disease when test is negative.
<sup>5</sup> Youden's J index is defined as sensitivity plus specificity minus 1 (perfect score = 1)
<sup>6</sup> F1 score is the harmonic mean of sensitivity and PPV (perfect score = 1)
<sup>7</sup> Difference in prevalence is the difference in the number of positive symptom screens (i.e. true positive plus false positive) for each combination from the actual number of RT-PCR positive specimens.
<sup>8</sup> Includes measured or subjective fever
<sup>9</sup> Included nasal congestion or runny nose.
<sup>10</sup> Cough, shortness of breath includes only if symptom was new or worsening in the 14 days prior to testing.

### Random forest analysis

Figure 1 shows the out-of-bag Receiver Operating Characteristic (ROC) curves for our five random forests, organized in ascending order by depth (*n*) and plotted against the full set of ROC points for the *m*-of-*n* combinations of equal size *n*. Depth did not substantially impact performance, with all forests achieving sensitivities of 69% and 74% at specificities of 95% and 90%, respectively. We can also see that the ROC curves generally contain the points for the combinations, suggesting that the performance of most combinations should generalize to new samples from the same population. Regardless of depth, the strongest predictors by feature importance were antigen test result, loss of taste/smell, chills, myalgia, and fever, many of which also appeared in the top-performing combinations.

**Figure 1.**
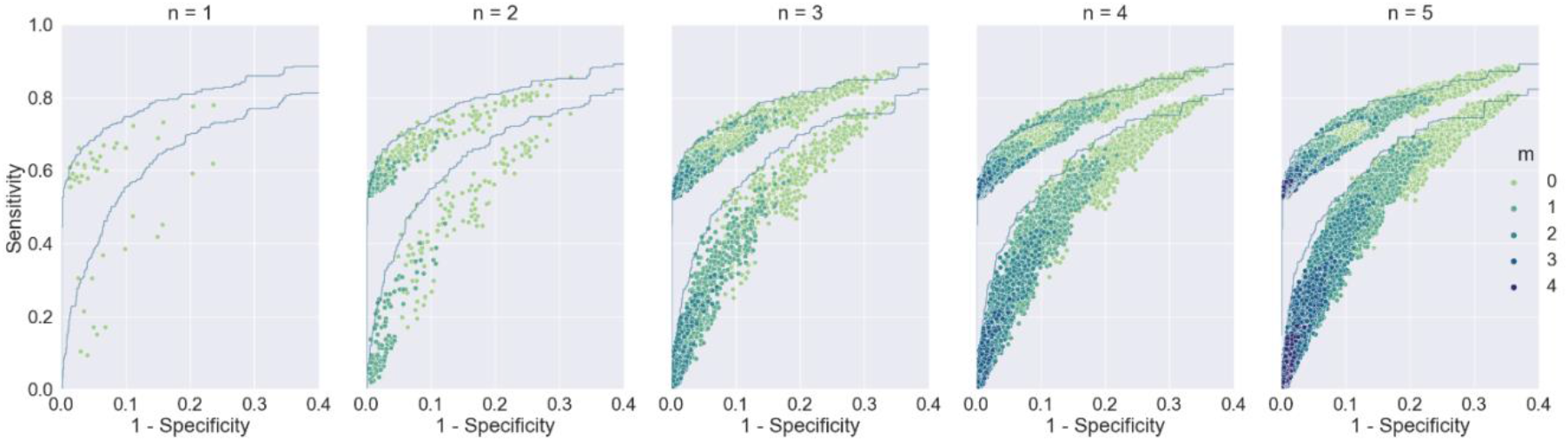
ROC curves for random forests of depth *n*, along with ROC points for *m*-of-*n* combinations of symptoms, both excluding (lower regions) and including (higher regions) antigen test results. Darker colors correspond to higher values of *m*, and each subplot specifies a different value for *n*. For both methods, including antigen test results improves sensitivity, and for the *m-*of-*n* combinations, increasing *m* increases specificity.

### Combinatorial analysis

Figure 1 also shows the enumeration of symptom combinations in ROC space. Higher values of *m* make the combinations less sensitive but more specific, and a lower ratio of *m* to *n* makes them more sensitive and less specific; increasing *n* has no substantial impact on performance alone. We see this trend in values of Youden’s J index, as well, which decreases as a function of *m* (average J was 0.50, 0.46, 0.38, 0.32, 0.28 for combinations with *m* of 1, 2, 3, 4, and 5, respectively) but not *n* (average J was 0.39 for all). By these measures, then, 1-of-*n* rules perform the best. Furthermore, because these rules are comparable to random forests of depth *n*, we expect those on or near the corresponding forest’s ROC curve both to generalize to new data and to be close to optimal.

The maximum achievable sensitivity for any combination of symptoms without antigen testing was 83% (specificity 61%); with antigen testing, this rose to 89% (specificity 61%). Because these are maxima at low values of *m* (1) and high values of *n* (15), and because they are well beyond the corresponding RF ROC curves, we do not expect them to generalize well to other datasets; nonetheless, they serve as useful points of comparison in selecting candidate rules we do expect to generalize.

For combinations near the ROC boundary, we propose three rules that seem to work particularly well in combination with antigen testing. In order of increasing specificity, these are as follows: one of fever, chills, fatigue, myalgia, or loss of taste or smell (82% sensitivity at 80% specificity); one of chills, difficulty breathing, or loss of taste or smell (75% at 91%); and two of rigors (shivers), headache, or loss of taste or smell (68% at 96%).

For ease of reference, we name these *spec80, spec90*, and *spec95*, respectively. Table 2 shows their classification performance both with and without antigen testing in comparison to loss of taste or smell alone and the existing case definitions (with the exception of the CSTE definition, none of these rules consider known exposure status).

**Table 2.**
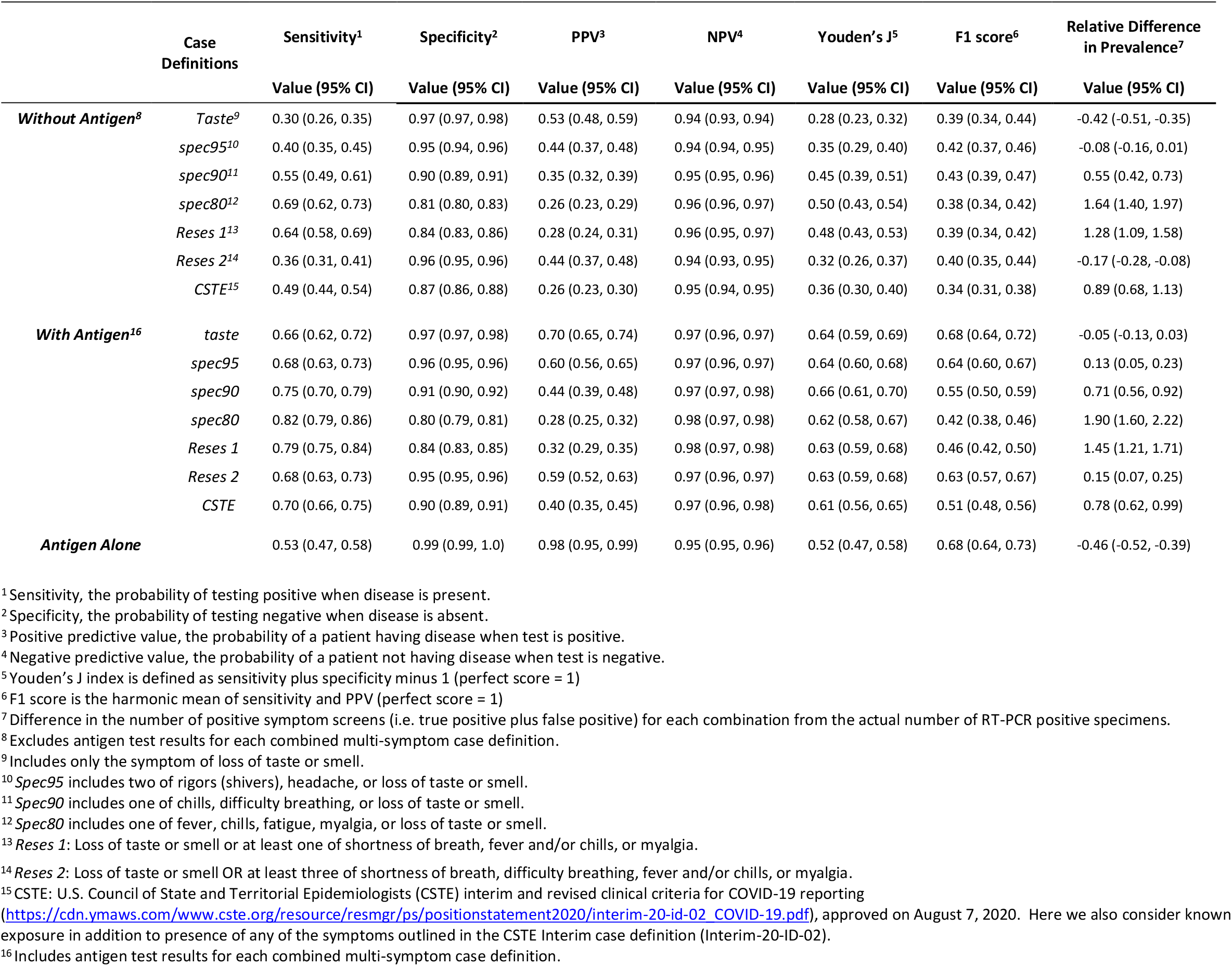
Point estimates and 95% bootstrap confidence intervals for the classification performance of multi-symptom case definitions with and without antigen test results. The CSTE definition additionally considers exposure status.

Of note, the CSTE case definition is the only one that does not perform optimally; all others, including *Reses 1* and *Reses 2*, achieve near-maximal levels of sensitivity at their given levels of specificity. This result is easier to see in Figure 2, which visualizes points for the case definitions against the full enumeration of *m*-of-*n* combinations in ROC space. While no definition is more specific than antigen testing alone, all are more sensitive, and some achieve a better balance of the two metrics, scoring up to 0.14 higher on J (0.52 for antigen alone versus 0.66 for *spec90*).

**Figure 2.**
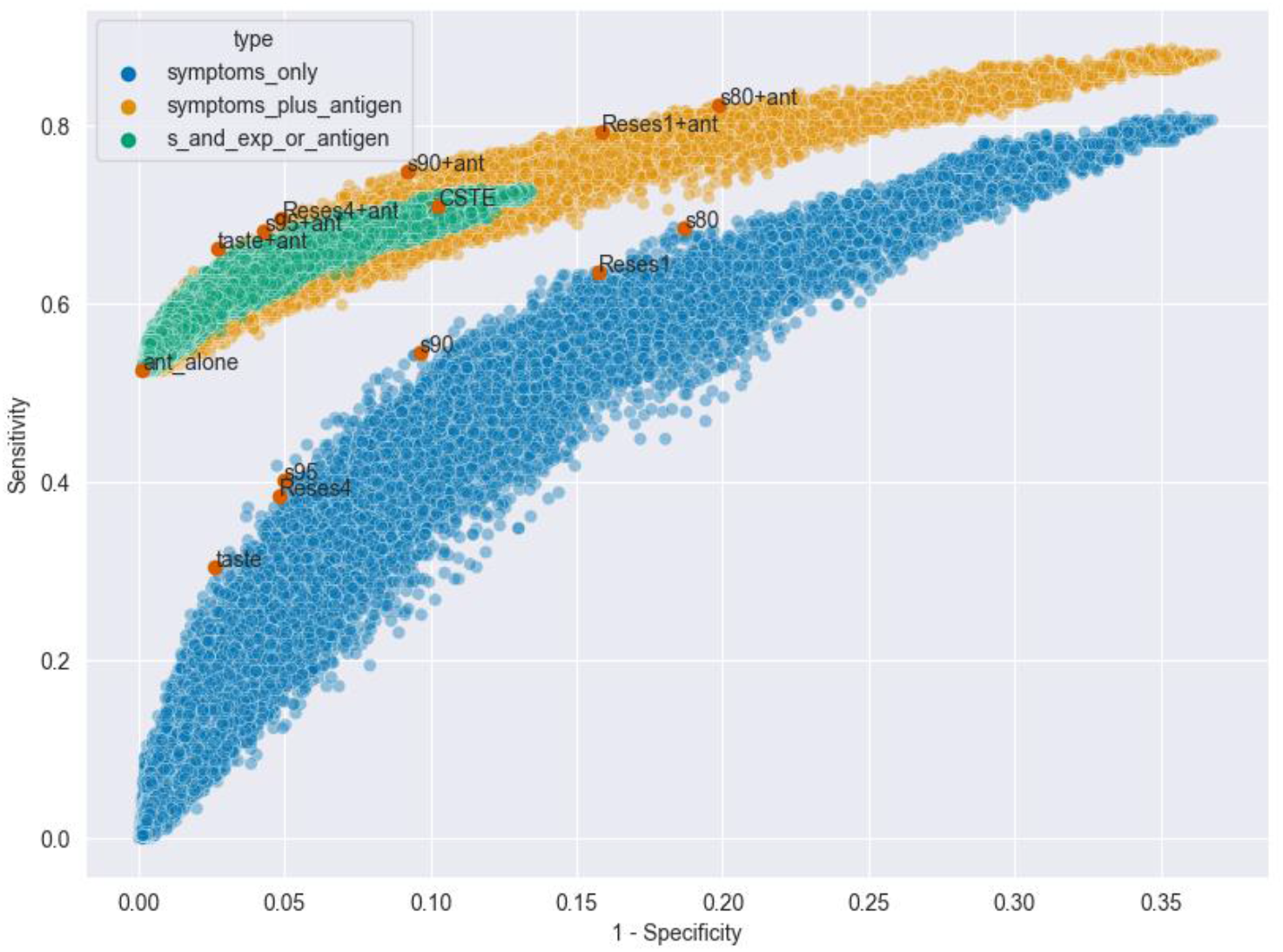
ROC points for preexisting and novel case definitions (red) against the set of points for the full enumeration of *m-*of-*n* combinations (blue, orange, and green). The colors correspond to the three kinds of predictors that could be included in a combination: symptoms alone (blue), symptoms plus antigen test results (orange), and symptoms plus antigen test results, but with the added requirement that symptoms are only considered with known exposure to a confirmed case (green).

For surveillance purposes, loss of taste or smell, when combined with antigen results, seems to be a good candidate, since it produces a nearly perfect prevalence estimate (−5%) in addition to achieving a relatively high score on J (0.64). This balance between group- and individual-level performance was rare in these data and suggests the rule could be used effectively for both screening and surveillance, especially considering its parsimony (only 2 features evaluated).

### The Benefit of Adding Exposure Status

Adding exposure status to the combinations generally did not improve their performance. When antigen test results were included alongside the symptoms, adding exposure status to a given *m*-of-*n* combination increased its J index only 43% of the time, lowering it the other 57% of the time (0.2% of the pairs had ties). Although the median J index was higher for rules considering exposure status (0.58) than for those not (0.57), the distribution of pairwise differences suggested the opposite (though very subtle) effect, with the median difference hovering around 0.01 in favor of the latter (Wilcoxon signed-rank test p-value < 0.001). Supplemental Figure 2 shows the distribution of these differences, and Supplemental Figure 3 shows how the distribution changes with respect to the specificity of the original combination (i.e., the combination without exposure status). Adding exposure status always made the original combination more specific, but when the original specificity was already high (more than approximately 0.80), the increase did not outweigh the resulting drop in sensitivity (Supplemental Figure 3).

## Discussion

In our case, adding reported symptom data to antigen test results boosted the latter’s sensitivity as a screening procedure without incurring a large drop in specificity. Given the basic characteristics of the data in our study, this is perhaps unsurprising: of the 142 study participants who were antigen negative but RT-PCR positive, 110 (78%) were symptomatic, and so the potential gain in sensitivity from considering symptoms was large from the start. Many of these missed cases are recouped by our proposed hybrid definitions (especially *spec80*), but the added sensitivity does come at the cost of reduced specificity, which under some practical circumstances may be non-negligible. This result highlights an important point for public health practice: case definitions should be tailored to their specific use cases, or, perhaps more precisely, to the real-world costs associated with false-positive and false-negative classifications.

From a methodological standpoint, our results suggest that using parsimony as a heuristic for selecting definitions may increase the chances of their performance generalizing to unseen data. *Reses1* and *Reses2* were both developed with a brute-force combinatorial search on different data (as rules with an *m*_*1*_-of-*n*_*1*_ OR *m*_*2*_*-*of-*n*_*2*_ structure, they are also more complex than the simple *m-*of-*n* definitions considered here) (17), and yet they performed well here, skirting the upper boundaries in ROC space for both the random forest and the full enumeration of symptom combinations. Although this kind of combinatorial search is the most straightforward way of determining which of the candidate combinations work best, it is computationally intensive and may be impractical to run on consumer hardware for very large feature spaces. In these scenarios, a hybrid approach may be sensible, such that a random forest or other machine learning method with a built-in variable-selection/ranking procedure (e.g., LASSO or L1-regularized regression) could be used to whittle down the feature space before performing the combinatorial search.

Relating to SARS-CoV-2 specifically, our results suggest that antigen testing alone may not always be the best strategy for screening or diagnosis in the absence of confirmatory testing (or even in the presence of confirmatory testing, which may vary depending on likelihood of SARS-CoV-2 infection and symptom status), which is in line with current CDC guidance on the use of antigen tests in the community (25). However, given the increase in use of antigen tests and the high demand for rapid test results (26), our analysis here shows the potential benefit for use of antigen tests in the context of specific symptoms to improve sensitivity, albeit at some loss of specificity. Screening scenarios where frequent, repeated antigen testing would be impractical—at border entry points where travelers or migrants may only be tested once before moving on, for example, or in private homes where the cost of repeat testing may be prohibitive—may be good settings for the use of hybrid screening rules that improve on the sensitivity of a single test. Conversely, when confirmatory tests are relatively scarce but required for care, or when the local healthcare system is nearing capacity, a more specific rule, like our *spec90* candidate rule or antigen testing alone, may be more appropriate. Finally, case definitions considering exposure status may be less optimal than those that do not, at least in scenarios similar to those in which our data were collected. In our results, these rules were often less sensitive than alternatives achieving the same specificity and less specific than those achieving the same sensitivity. This effect may have been due to the presence of other high-performing predictors in the data, like the antigen test results and loss of taste and/or smell, that made the added specificity from considering exposure less useful than it would have been otherwise.

Our analysis has several limitations. First, the CSTE COVID-19 case definition (27) has been updated since the time of this investigation to add additional symptoms for consideration which were not included in our investigation and cannot be evaluated, although many symptoms added to the most recent definition are more severe in nature (confusion or change in mental status; persistent chest pain/pressure; blue-colored skin, lips, or nail beds, depending on skin tone; and inability to wake or stay awake), and it is unlikely they would have been captured in an outpatient community setting. Second, although SARS-CoV-2 samples were not sequenced as part of this investigation, data collection occurred in November 2020, well before the two variants of concern, Delta (B.1.617.2) and Omicron (B.1.1.529), were documented in the United States. The candidate definitions we propose above, which rely heavily on loss of taste or smell to boost the sensitivity of antigen testing, may not generalize well to populations heavily affected by these variants if the symptoms they cause differ substantially from those cause by the original strain. Additionally, it is unclear whether these results may apply to re- infections or vaccine-breakthrough infections as this study was conducted prior to vaccine availability and widespread concern for re-infection. Because our analysis could easily be rerun with the availability of new data, we hope these limitations will be addressed by future research. Third, the data presented here are cross-sectional and pre-symptomatic individuals could not be identified. Fourth, as mitigation measures have relaxed, such as limits on large gatherings, mask use, etc., the circulation of other non-SARS-CoV-2 respiratory viruses will likely increase and the impact on symptom analysis for SARS-CoV-2 is not fully known. Finally, the FDA recently published a statement (28) suggesting that antigen tests may have reduced sensitivity against the Omicron variant, although there is also some recent data showing antigen test sensitivity remains similar to pre-Omicron levels in a community setting (29).

## Conclusion

While confirmatory testing for symptomatic persons with a negative antigen test is recommended (25), this often presents challenges including logistical and cost constraints and longer turnaround times for RT-PCR, thus delaying care and/or isolation of individuals (26). In certain situations, confirmatory testing may not be available at all – especially in low resource settings. Therefore, using the methodology described in this study to better understand combinations of symptoms that are more predictive of SARS-CoV-2 infection in the absence of a positive antigen test may aid clinicians and public health professionals. In situations where confirmatory testing is not available, and immediate public health action is needed, considering the presenting symptoms in antigen negative individuals may provide important information on how to prioritize public health action, especially in congregate living or school settings.

## Data Availability

All data in the present study are available upon reasonable request provided to the authors.

## DISCLAIMER

The findings and conclusions in this report are those of the authors and do not necessarily represent the official positions of the Centers for Disease Control and Prevention (CDC).

## References

1. Centers for Disease Control and Prevention. COVID Data Tracker. Trends in Number of COVID-19 Cases and Deaths in the US Reported to CDC, by State/Territory. Atlanta, GA: US Department of Health and Human Services, CDC; 2022, March 24. https://covid.cdc.gov/covid-data-tracker/#trends_dailycases,

2. Pilarowski G, Marquez C, Rubio L, et al. Field performance and public health response using the BinaxNOW TM Rapid SARS-CoV-2 antigen detection assay during community-based testing. Clin Infect Dis 2020.

3. Almendares O, Prince-Guerra JL, Nolen LD, et al. Performance Characteristics of the Abbott BinaxNOW SARS-CoV-2 Antigen Test in Comparison to Real-Time Reverse Transcriptase PCR and Viral Culture in Community Testing Sites during November 2020. J Clin Microbiol 2022;60(1):e0174221.

4. McKay SL, Tobolowsky FA, Moritz ED, et al. Performance Evaluation of Serial SARS-CoV-2 Rapid Antigen Testing During a Nursing Home Outbreak. Ann Intern Med 2021.

5. James AE, Gulley T, Kothari A, et al. Performance of the BinaxNOW coronavirus disease 2019 (COVID-19) Antigen Card test relative to the severe acute respiratory coronavirus virus 2 (SARS-CoV-2) real-time reverse transcriptase polymerase chain reaction (rRT-PCR) assay among symptomatic and asymptomatic healthcare employees. Infect Control Hosp Epidemiol 2021:1–3.

6. Pollock NR, Jacobs JR, Tran K, et al. Performance and Implementation Evaluation of the Abbott BinaxNOW Rapid Antigen Test in a High-Throughput Drive-Through Community Testing Site in Massachusetts. J Clin Microbiol 2021;59(5).

7. Shah MM, Salvatore PP, Ford L, et al. Performance of Repeat BinaxNOW SARS-CoV-2 Antigen Testing in a Community Setting, Wisconsin, November-December 2020. Clin Infect Dis 2021.

8. Sood N, Shetgiri R, Rodriguez A, et al. Evaluation of the Abbott BinaxNOW rapid antigen test for SARS-CoV-2 infection in children: Implications for screening in a school setting. PLoS One 2021;16(4):e0249710.

9. Allan-Blitz LT, Klausner JD. A Real-World Comparison of SARS-CoV-2 Rapid Antigen Testing versus PCR Testing in Florida. J Clin Microbiol 2021;59(10):e0110721.

10. Prince-Guerra JL, Almendares O, Nolen LD, et al. Evaluation of Abbott BinaxNOW Rapid Antigen Test for SARS-CoV-2 Infection at Two Community-Based Testing Sites -Pima County, Arizona, November 3-17, 2020. MMWR Morb Mortal Wkly Rep 2021;70(3):100–5.

11. Okoye NC, Barker AP, Curtis K, et al. Performance Characteristics of BinaxNOW COVID-19 Antigen Card for Screening Asymptomatic Individuals in a University Setting. J Clin Microbiol 2021;59(4).

12. Surasi K, Cummings KJ, Hanson C, et al. Effectiveness of Abbott BinaxNOW Rapid Antigen Test for Detection of SARS-CoV-2 Infections in Outbreak among Horse Racetrack Workers, California, USA. Emerg Infect Dis 2021;27(11):2761–7.

13. Ford L, Lee C, Pray IW, et al. Epidemiologic characteristics associated with SARS-CoV-2 antigen-based test results, rRT-PCR cycle threshold values, subgenomic RNA, and viral culture results from university testing. Clin Infect Dis 2021.

14. Frediani JK, Levy JM, Rao A, et al. Multidisciplinary assessment of the Abbott BinaxNOW SARS-CoV-2 point-of-care antigen test in the context of emerging viral variants and self-administration. Sci Rep 2021;11(1):14604.

15. U.S. Council for State and Territorial Epidemiologists (CSTE). Coronavirus Disease 2019 (COVID-19) 2020 Interim Case Definition, Approved August 5, 2020: CSTE Position Statement. https://cdn.ymaws.com/www.cste.org/resource/resmgr/ps/positionstatement2020/interim-20-id-02_COVID-19.pdf, 2020, mAugust 5,

16. Cain KP, McCarthy KD, Heilig CM, et al. An algorithm for tuberculosis screening and diagnosis in people with HIV. N Engl J Med 2010;362(8):707–16.

17. Reses HE, Fajans M, Lee SH, et al. Performance of existing and novel surveillance case definitions for COVID-19 in household contacts of PCR-confirmed COVID-19. BMC Public Health 2021;21(1):1747.

18. See e.g. 45 C.F.R. part 46.102(l)(2), 21 C.F.R. part 56; 42 U.S.C. Sect. 241(d); 5 U.S.C. Sect. 552a; 44 U.S.C. Sect. 3501 et seq. https://www.hhs.gov/ohrp/sites/default/files/ohrp/policy/ohrpregulations.pdf,

19. U.S. Food and Drug Administration. CDC 2019-Novel Coronavirus (2019-nCoV) Real-Time RT-PCR Diagnostic Panel - Instructions for Use. https://www.fda.gov/media/134922/download, 2020,

20. Fosun Pharma USA. Instruction for Use: Fosun COVID-19 RT-PCR Detection Kit. https://www.fda.gov/media/137120/download, 2020,

21. Breiman L. Random Forests. Machine Learning 2001;45(1):5–32.

22. Efron B. Better Bootstrap Confidence Intervals. Journal of the American Statistical Association 1987 Mar;82(397):171–85.

23. Pedregosa F VG, Gramfort A, Michel V, Thirion B, Grisel O, Blondel M, Prettenhofer P, Weiss R, Dubourg V, Vanderplas J. Scikit-learn: Machine learning in Python. The Journal of machine Learning research 2011 Nov 1;12:2825–30.

24. Seabold S, Perktold J. Statsmodels: Econometric and Statistical Modeling with Python. 2010.

25. Centers for Disease Control and Prevention. Coronavirus Disease 2019 (COVID-19): Interim Guidance for Antigen Testing for SARS-CoV-2. Atlanta, GA: US Department of Health and Human Services, CDC; 2021. https://www.cdc.gov/coronavirus/2019-ncov/lab/resources/antigen-tests-guidelines.html, 2022,

26. Peeling RW, Olliaro PL, Boeras DI, et al. Scaling up COVID-19 rapid antigen tests: promises and challenges. Lancet Infect Dis 2021;21(9):e290–e5.

27. U.S. Council for State and Territorial Epidemiologists (CSTE). Coronavirus Disease 2019 (COVID-19) 2021 Case Definition. https://ndc.services.cdc.gov/case-definitions/coronavirus-disease-2019-2021/, 2021,

28. FDA. SARS-CoV-2 Viral Mutations: Impacts on COVID-19 Tests. US Department of Health and Human Services. https://www.fda.gov/medical-devices/coronavirus-covid-19-and-medical-devices/sars-cov-2-viral-mutations-impact-covid-19-tests#omicronvariantimpact, 2021.,

29. Schrom J, Marquez C, Pilarowski G, et al. Direct Comparison of SARS-CoV-2 Nasal RT-PCR and Rapid Antigen Test (BinaxNOW™) at a Community Testing Site During an Omicron Surge. medRxiv 2022:2022.01.08.22268954.

