## Supplemental Figures for "Performance of Existing and Novel Symptom- and Antigen Testing-Based COVID-19 Case Definitions in a Community Setting"

**Supplementary Data**

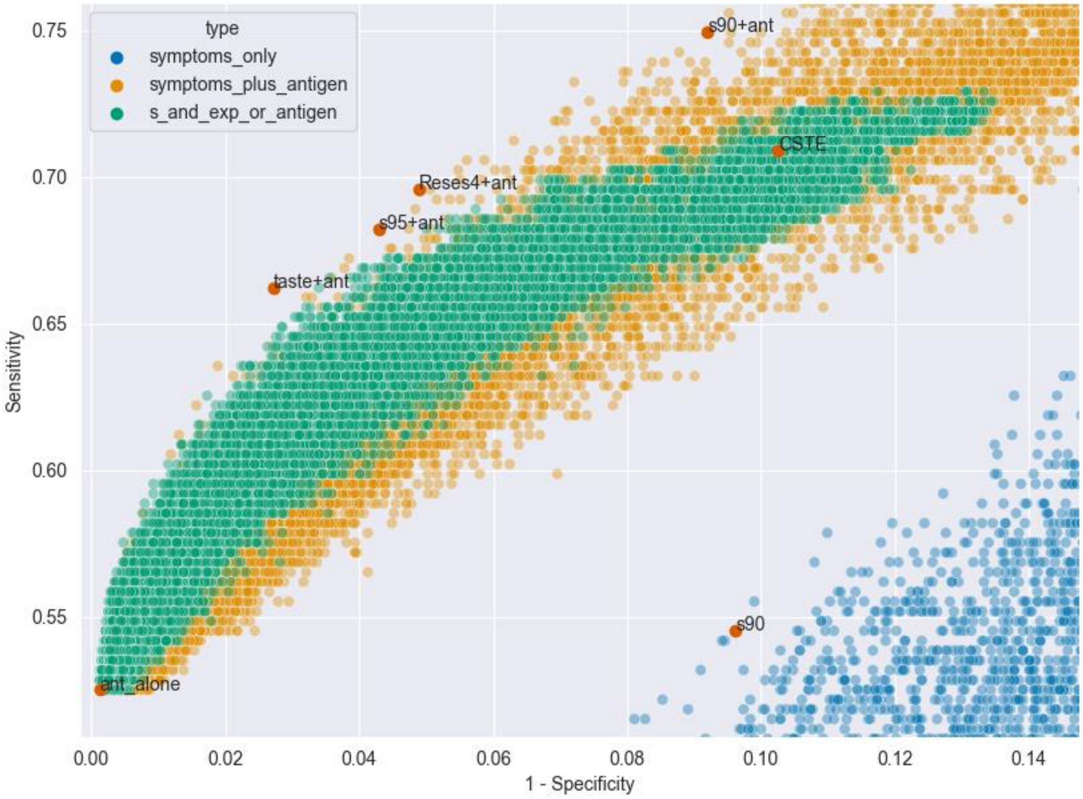


**Supplemental Figure 1.** Case definitions including antigen test results in ROC space. Definitions not considering exposure status (yellow) begin to outperform those that do (green) at a specificity of around 95%. The CSTE definition, which considers exposure status and contains a compound symptom criterion (i.e., either two of a set of more general symptoms or one of a set of COVID-specific symptoms), is more sensitive than about half of definitions achieving the same specificity.

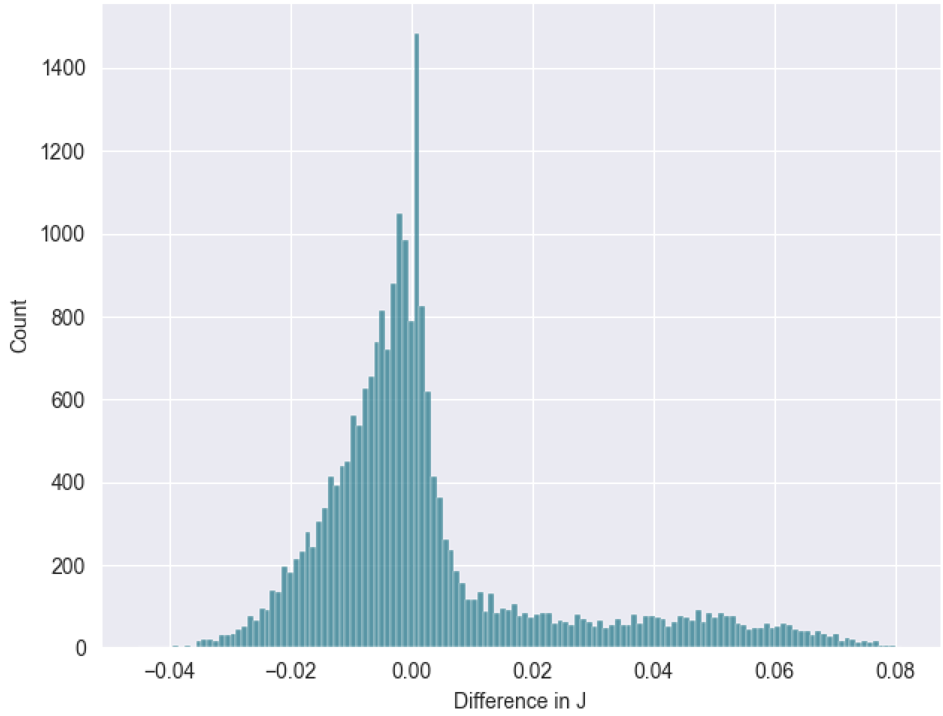


**Supplemental Figure 2**. The distribution in pairwise differences in overall classification performance as measured by J Index between case definitions with and without exposure status included.

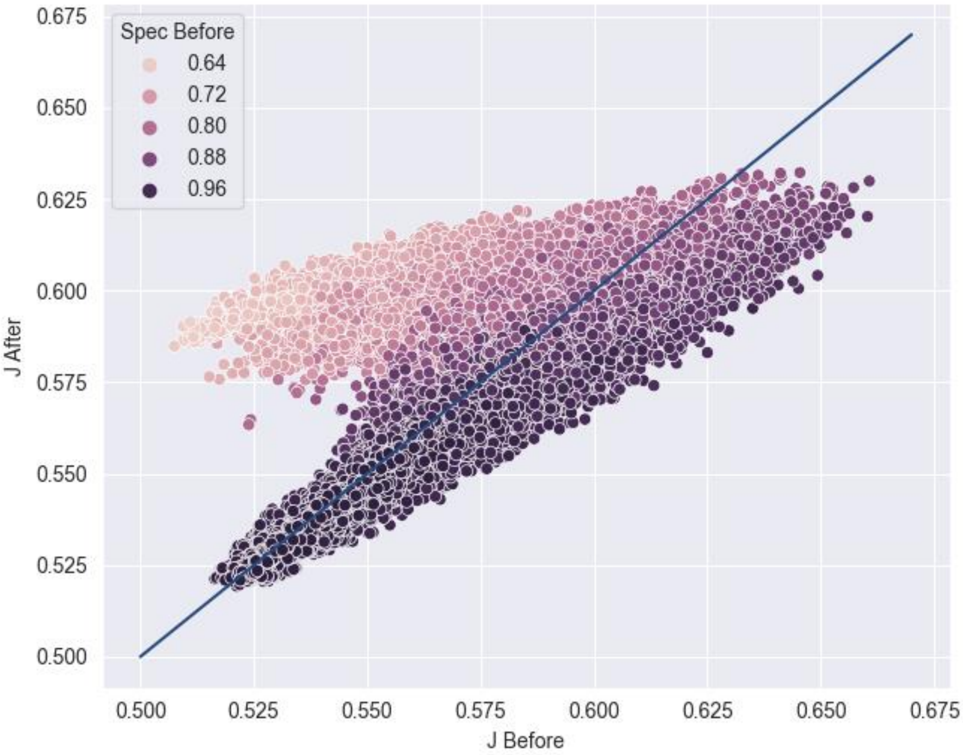


**Supplemental Figure 3.** The relationship between overall classification performance before (x-axis) and after (y-axis) adding exposure status to a case definition, with the zero-change line running on the diagonal. Definitions benefited more from including exposure status when their original specificity was low. All definitions included antigen test results and symptoms to start.
